# An economic evaluation of two cervical screening algorithms in Belgium: HR-HPV primary compared to HR-HPV and liquid-based cytology (LBC) co-testing

**DOI:** 10.1101/2023.09.07.23295193

**Authors:** Caroline Dombrowski, Claire Bourgain, Yixuan Ma, Anne Meiwald, Amy Pinsent, Birgit Weynand, Katy ME Turner, Susie Huntington, Elisabeth J Adams, Johannes Bogers, Romaric Croes, Shaira Sahebali

## Abstract

**Objective:** To assess the costs and benefits of two algorithms for cervical cancer screening in Belgium 1) high-risk human papillomavirus (HR-HPV) primary screening and 2) HR-HPV and liquid-based cytology (LBC) co-testing.

**Methods:** A decision tree was adapted from published work and parameterised using HORIZON study data and Belgian cost and population data. The theoretical model represents two different screening algorithms for a cohort of 577,846 women aged 25 – 64 attending routine cervical screening. Scenario analyses were used to explore the impact of including vaccinated women and alternative pricing approaches. Uncertainty analyses were conducted.

**Results:** The cost per woman screened was €113.50 for HR-HPV primary screening and €101.70 for co-testing, representing a total cost of €65,588,573 and €58,775,083 respectively for the cohort; a 10% difference. For one screening cycle, compared to HR-HPV primary, co-testing resulted in 13,173 more colposcopies, 67,731 more HR-HPV tests and 477,020 more LBC tests. Co-testing identified 2,351 more CIN2+ cases per year (27% more than HR-HPV primary) and 1,602 more CIN3+ cases (24% more than HR-HPV primary) than HR-HPV primary.

**Conclusion:** In Belgium, a co-testing testing algorithm could increase cervical precancer detection rates compared to HR-HPV primary. Co-testing would cost less than HR-HPV primary if the cost of the HPV test and LBC were cost-neutral compared to the current cost of LBC screening but would cost more if the cost per HPV test and LBC were the same in both co-testing and HR-HPV primary strategies.

## Introduction

Cervical cancer, caused by high-risk human papillomavirus (HR-HPV), is a leading cause of mortality in women (WHO, 2022). While not all HR-HPV infections result in cancer, persistent HR-HPV infection can lead to cervical abnormalities, which if not detected and treated, progress to invasive cervical cancer (Monsonego et al., 2004).

The World Health Organization (WHO) has set a global target to eliminate cervical cancer through a combined approach of prevention (HR-HPV vaccination), testing (cervical screening programmes) and treatment (WHO, 2020a). Cervical screening can identify HR-HPV and cervical cell abnormalities that, when detected at an early stage, can be managed to reduce the risk of disease progression and mortality (Jansen et al., 2020). While many high-income countries have offered high-quality cervical screening via Pap smear tests for many years, technological advancements in testing and interventions, including HR-HPV vaccination, have led countries to review their existing programmes (Anttila et al., 2004). Studies demonstrated the benefits of HPV-based screening (Ronco et al., 2014) and some countries have or will transition from cytology-based screening to HR-HPV primary testing with LBC (liquid-based cytology) following an HR-HPV positive result (Maver and Poljak, 2020)). Other countries, like the United States (US) and Germany (Fontham et al., 2020; Xhaja et al., 2022), have chosen to implement co-testing (using LBC and HR-HPV testing concurrently) as studies suggest that co-testing can detect more people who are HR-HPV negative but have abnormal cytology. Approximately 3-14% (precancer) or 13-18% (cancer) cases are HPV negative and are therefore missed using HR-HPV primary testing (Austin R.M. et al., 2018; Vasilyeva et al., 2021, Blatt et al., 2015a; Kaufman et al., 2020). F

Data from a large study in Belgium in 2010 indicates the prevalence of HR-HPV is around 13% in women (Depuydt et al., 2010), with approximately 3,000 cases of carcinoma *in situ* (Arbyn Marc et al., 2015) and 600 new cases of invasive cervical cancer diagnosed annually (Bruni et al., 2023). Although access to screening is available, there is currently no centrally organised national cervical screening programme. Screening coverage was reported as 61% nationally for the period 2004-2006 (Arbyn et al., 2014) and more recently, as 62.6% in Flanders (Flemish Department of care, 2021). Regional health authorities are responsible for screening and their approaches vary. In Flanders, women aged 25 – 64 receive an LBC test every three years in a semi-organised screening programme, while in French-speaking regions, screening is opportunistic (Van Kerrebroeck and Makar, 2016). In 2020, a pilot screening program for women aged 25-64 launched in Wallonia. HR-HPV vaccination coverage varies considerably by region from 91% in Flanders (Thiry et al., 2019) to 31-50% in French-speaking regions (Bonanni et al., 2020).

As the Belgium government considers moving from a regionally organised cytology-based screening to a national HR-HPV primary screening programme, it is important to consider the costs and potential benefits of different screening algorithms. Health economic models can be used to inform decision making by quantifying the outcomes of different screening algorithms within a given setting (Mendes et al., 2015). This study uses a decision tree model to assess the health and economic outcomes of a HR-HPV primary screening algorithm compared to co-testing algorithm in Belgium.

## Methods

### Model overview

A decision tree model was developed in Excel v2202 (Microsoft, Redmond, WA, USA) to simulate potential cervical screening algorithms for Belgium, comparing HR-HPV primary with co-testing algorithms. A cost-consequence analysis was undertaken from the perspective of the Belgian healthcare system.

### Cervical screening algorithm

A national cervical screening algorithm for HR-HPV primary screening in Belgium has not yet been finalised, therefore theoretical cervical screening algorithms were developed for the Belgium setting (**Figure 1**); the HR-HPV primary algorithm was adapted from the Netherlands screening algorithm (RIVM, 2021) and the co-testing algorithm was adapted from screening algorithms in Germany and Luxembourg (Iftner, 2021; Secrétariat du Conseil Scientifique, 2019).

**Figure 1.**
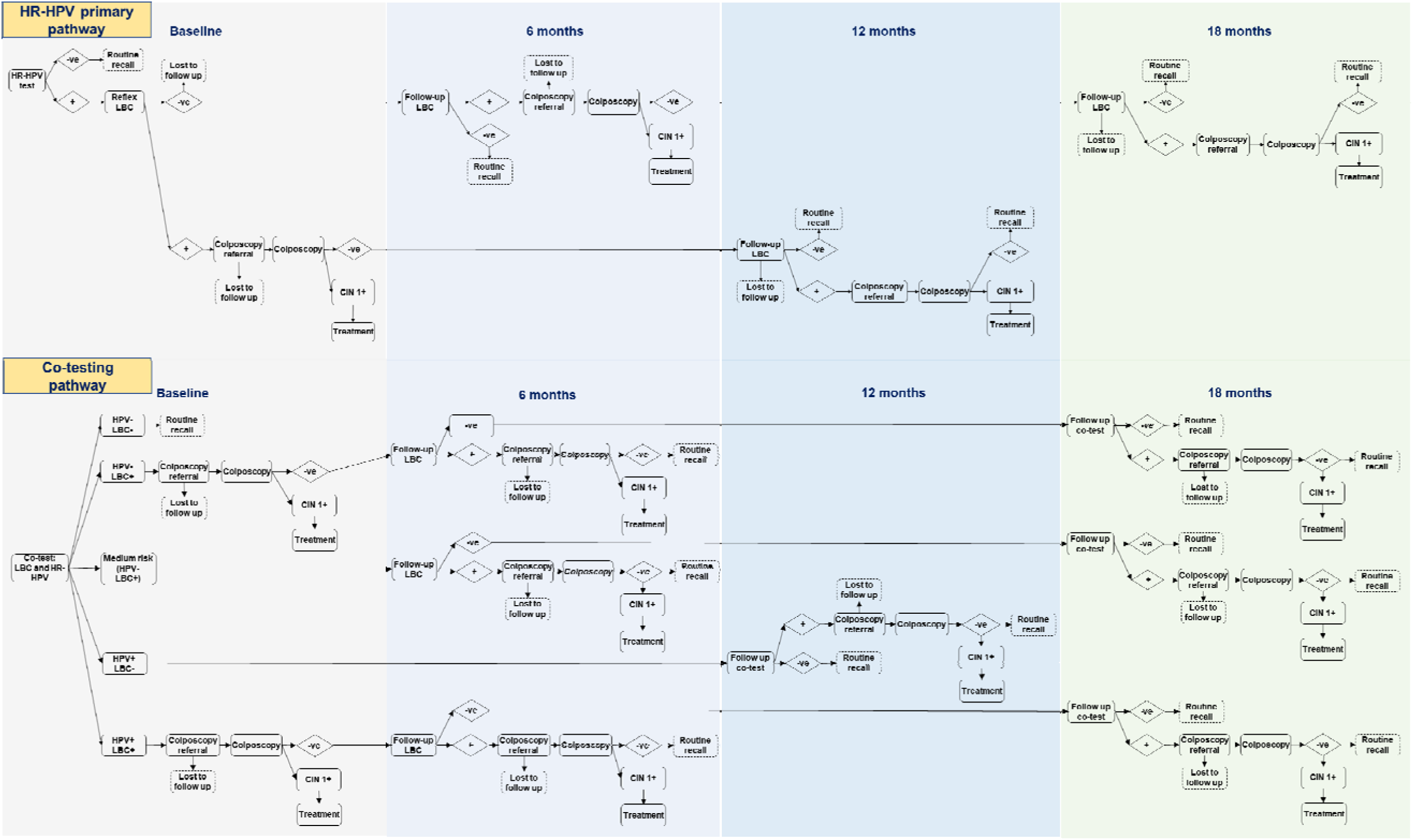
HR-HPV primary and co-testing screening pathways.

In the HR-HPV primary algorithm, women have a cervical sample taken which is tested for HR-HPV (**Figure 1**). Those who test negative return for re-testing every 5 years (hereafter referred to as the routine screening cycle) and HR-HPV positive samples are tested using reflex-LBC Those with normal baseline LBC results return after 6 months for repeat LBC testing. Those with abnormal (non-NILM) LBC results are referred for colposcopy. If the colposcopy and histology results are abnormal, they receive appropriate treatment, or if the results are normal, they return for a follow-up LBC in 12 months.

In the co-testing algorithm, all cervical samples have HR-HPV testing and LBC simultaneously (**Figure 1**). Those with HR-HPV negative and normal LBC results return to the routine screening cycle (5 years). Those with HR-HPV positive results and normal LBC results have a repeat co-test after 12 months and those with repeat HR-HPV positive and/or LBC abnormal results are referred for colposcopy, whereas those with negative results return to routine recall. At routine testing, those with abnormal LBC results are referred for colposcopy immediately and if the biopsy is negative, return for an additional LBC after 6 months; those with normal LBC results return for repeat co-testing after 18 months.

Like the German co-testing algorithm, the co-testing algorithm includes a medium-risk group to simulate a more conservative follow-up of women who are HR-HPV negative and have borderline or low-grade LBC results. Instead of direct referral to colposcopy, a proportion of those with HR-HPV negative and abnormal but low-grade LBC baseline results receive follow-up LBC testing at 6 months (**Table 1**), and if LBC results are normal, return for repeat co-testing at 18 months. Those with abnormal LBC results are referred to colposcopy. The remainder are assumed to have higher grade LBC results and are directly referred to colposcopy. Patients exit the algorithm to return to routine recall after negative co-testing results, after the second colposcopy with negative biopsy results, or after treatment after the biopsy. All samples were assumed to give conclusive test results, with no inadequate samples.

**Table 1.**
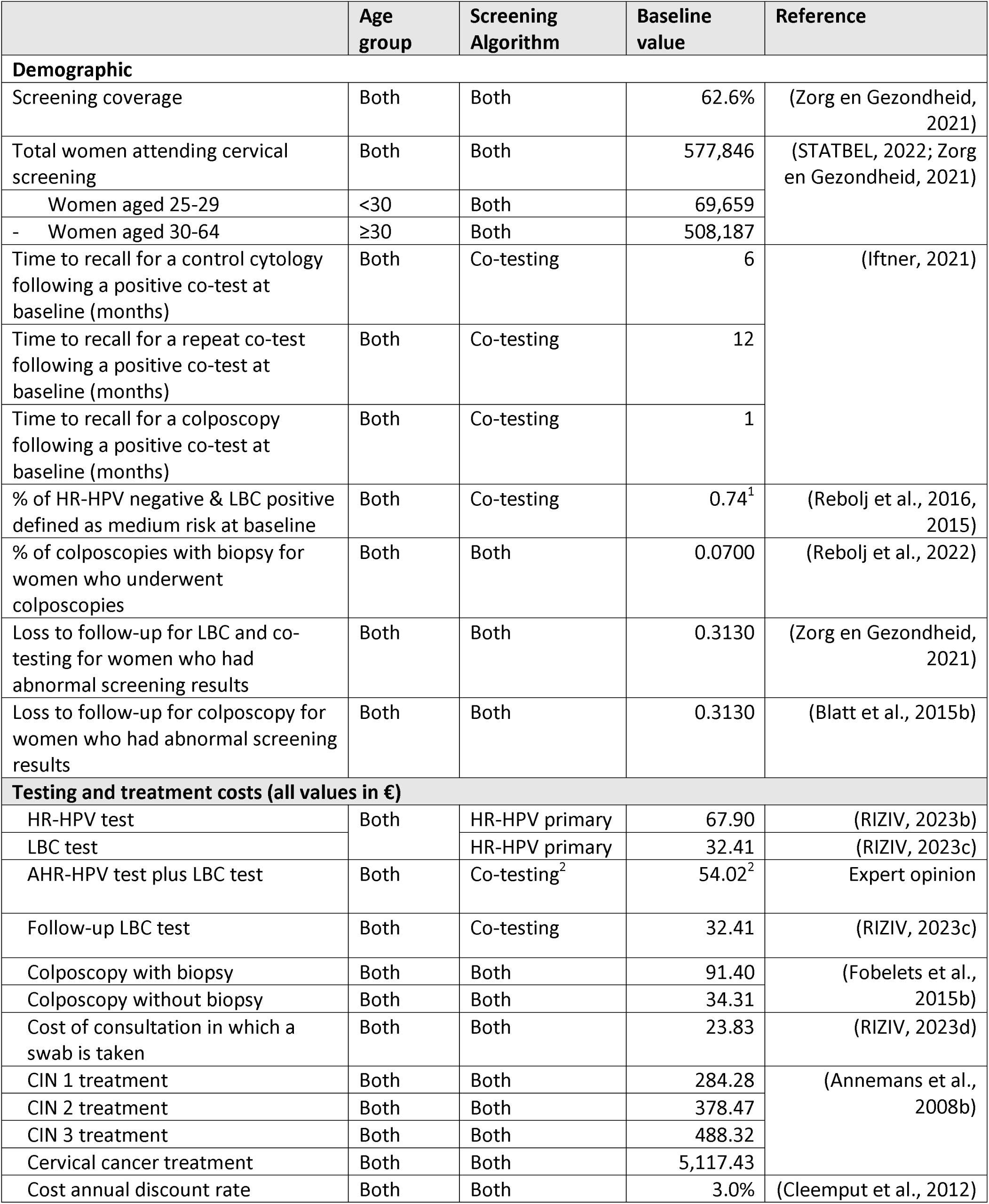
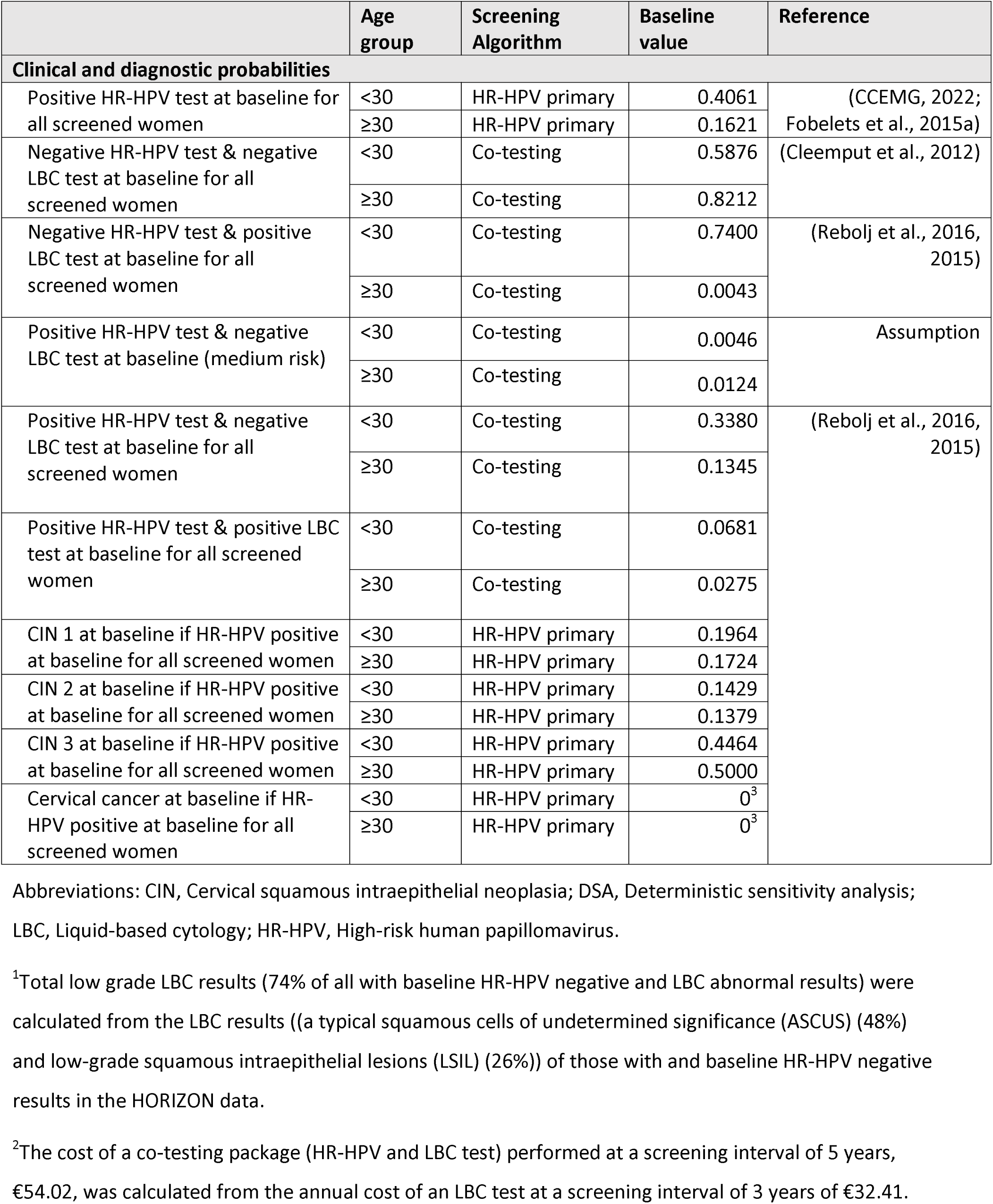
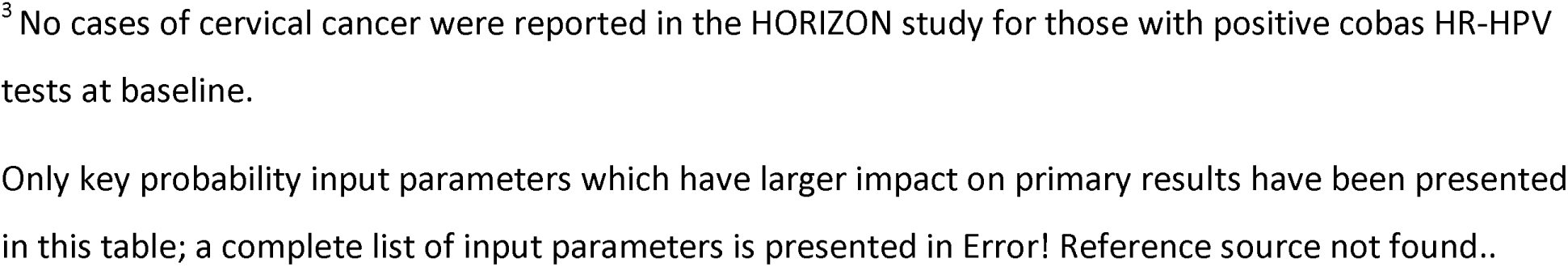
Key model input parameters baseline values.

### Patient and public involvement

This is a theoretical analysis, using only published and routinely available data. No patients were involved at any stage of this study and only published aggregate patient data was used.

### Outcomes

The primary outcomes of the study were total screening costs (from attendance at screening to treatment for cervical intraepithelial neoplasia (CIN1-CIN3)), average cost per complete screen, cost per CIN2+ and CIN3+ case, total number of colposcopies, number of HR-HPV and LBC tests performed, and number of cervical precancer and cancer cases diagnosed.

The secondary outcomes were the total cost of colposcopies, total cost of HR-HPV and LBC tests, and cost of treatment of CIN1, CIN2 and CIN3. The difference in costs and outcomes between the two screening algorithms were calculated (HR-HPV primary and co-testing).

### Population

The simulated population represents 577,846 women in Belgium aged 25-64 attending screening in one year, assuming a 5-year recall period. The most inclusive language for cervical screening is “women and people with a cervix” (Gov.uk, 2022); the term ‘women’ is used throughout this paper to reflect EU screening guidelines (European Commission, 2022) and Belgian literature (Arbyn Marc et al., 2015).

### Model parameters

The model parameter inputs are detailed in **Table 1** and Error! Reference source not found.**Supplemental Table 1** and include screening coverage, probability of detecting HR-HPV and pre-cancer (by age group), testing probabilities (HR-HPV primary, cytology, colposcopy), loss to follow up and cost inputs.

#### Transition probability inputs

There is limited published individual patient-level data reporting long-term health outcomes associated with different testing algorithms and no Belgian-specific data. The HORIZON study (Rebolj et al., 2016, 2015) (see **Supplemental Tables** for details), was conducted in Denmark and assessed 4,128 cervical samples using four different HR-HPV assays and LBC, was selected as an appropriate source to inform the clinical and diagnostic probability parameters.

The probability of detecting LBC abnormalities and abnormal colposcopy results (CIN1, CIN2 and CIN3) or worse (**Table 1**) were calculated using data from the HORIZON study based on the results for cobas 4800 HR-HPV assay (Rebolj et al., 2016, 2015). Methods for calculating the transition probabilities for primary HR-HPV testing are described in detail in a previous model (Weston et al., 2020). HORIZON reported one case of cervical cancer in the study population. Due to the small sample size, this was deemed not suitable for estimating cervical cancer in an entire screening population and therefore CIN3 or worse (CIN 3 and cervical cancer, CIN3+) was selected as the most appropriate outcome.

Screening coverage (62.6% baseline value) was informed using Flemish screening data (Flemish Department of care, 2021).

#### Cost inputs

Costs in Belgium were estimated using published data (Annemans et al., 2008a; Fobelets et al., 2015a; RIZIV, 2023a, 2023b, 2023c). All costs (presented in Euros) were inflated to 2023 values (**Table 1**) using the IMF inflation index (CCEMG, 2022).The cost of all tests includes sampling and processing costs. The cost of colposcopies and treatment were informed using published literature (Annemans et al., 2008b; Fobelets et al., 2015b). The cost of the initial consultation was taken from RIZIV, a Belgian government agency (RIZIV, 2023a). Costs incurred more than one year in the future were discounted by 3.0%, as recommended by Belgian Healthcare Knowledge Centre (KCE) (Cleemput et al., 2012).

As test reimbursement remains uncertain and may vary in different programs and settings, a cost-neutral pricing strategy was developed for co-testing in consultation with experts and is proposed as a potential pricing approach for Belgium. The cost of LBC (€32.41) in the current LBC-based screening algorithm which has a 3-year screening interval is equivalent to €10.80 per year. This yearly cost was used to calculate a cost-neutral price for the co-testing package (HR-HPV and LBC) which has a 5-year screening interval (i.e., €10.80 × 5 = €54.02). The discounting applied when HR-HPV testing and LBC are used for all samples (co-testing) compared to when HR-HPV testing is used for all samples, but LBC is used only for some (HR-HPV primary), is a commercial approach used in the past where two diagnostic tests are used together.

### Uncertainty analyses

#### Deterministic sensitivity analyses

Deterministic one-way sensitivity analyses (DSA) were conducted to assess the impact of changing each parameter on the main outcomes. Low and high parameter values were individually varied to generate results (**Supplemental Table 2**). The cost of colposcopy and treatment were varied by 20% to explore potential future variations in costs. The co-testing package cost (co-testing algorithm) was varied by €20; the cost of the HR-HPV test was varied by €20 and LBC by €10 in the HPV-primary algorithm.

Probabilities were varied based on the calculated standard error (SE). Those parameters with no SE value assigned were varied by +/− 25% of their baseline values. The low value for loss to follow-up was assumed to be zero, indicating 100% compliance. Tornado plots are used to present the DSA for overall costs, number of colposcopies, HR-HPV, LBC tests and cervical precancer and cancer diagnoses (**Error! Reference source not found. Figure 1**). Parameters that impacted the results by ≥10% are reported.

#### Probabilistic sensitivity analysis

Probabilistic sensitivity analysis (PSA) was performed to explore the robustness of results by running the model 1,000 times independently using a Monte Carlo simulation. All input cost parameters were assigned a Gamma distribution, and beta distribution was used for probability inputs from 0 to 1, which were calculated based on estimated alpha and beta values (**Supplemental Table 2, Supplemental Table 3**). The same values were used for parameters used in both co-testing and HR-HPV primary algorithms (e.g., probability of loss to follow-up for colposcopy). The 95% confidence intervals (CI) were calculated from the results of 1,000 iterations.

#### Scenario analyses

In the first two scenario analyses, an increased HR-HPV vaccination coverage was assumed (compared to 0% in the base case) for women <30 years to simulate HPV vaccinated women entering the screening cohort. The reduced probability of an HR-HPV positive test was calculated using relative risk data from published studies (**Supplemental Table 4**). Scenario 1 represented current vaccination coverage: 67% (Bruni et al., 2023), and Scenario 2, the target coverage: 90% (WHO, 2020b). In Scenario 3, the current reimbursement cost for the HR-HPV test (€67.90) (RIZIV, 2023c) and the LBC cost (€32.41) was used in both screening algorithms.

## Results

### Baseline results

The primary outcomes are presented in **Table 2**. Using the cost-neutral pricing strategy for co-testing, implementing co-testing, which has a different follow-up algorithm (**Figure 1**), would save an average of €11.80 per person screened compared to the HR-HPV primary algorithm. The co-testing algorithm would require an additional 13,173 colposcopies, 67,731 HR-HPV tests and 477,020 LBC tests for the screening cohort (the total programme costs of €58.8M for co-testing and €65.6M for HR-HPV primary in **Table 2**). However, co-testing, would detect an additional 2,351 CIN2+ cases (**Figure 2**), of which 1,602 would be CIN3+ cases, representing an increase of 27.4% and 24.3% respectively. The average cost per CIN2+ case detected was €5,381 for co-testing compared to €7,650 for HR-HPV primary (**Table 4**). Cost outcomes are presented in **Table 3**. The cost of HR-HPV testing contributed the largest component of the total cost in both screening algorithms (81% of total costs in HR-HPV primary and 56% for co-testing) (**Table 3**). The cost of LBC was the second largest contributor to overall costs.

**Figure 2.**
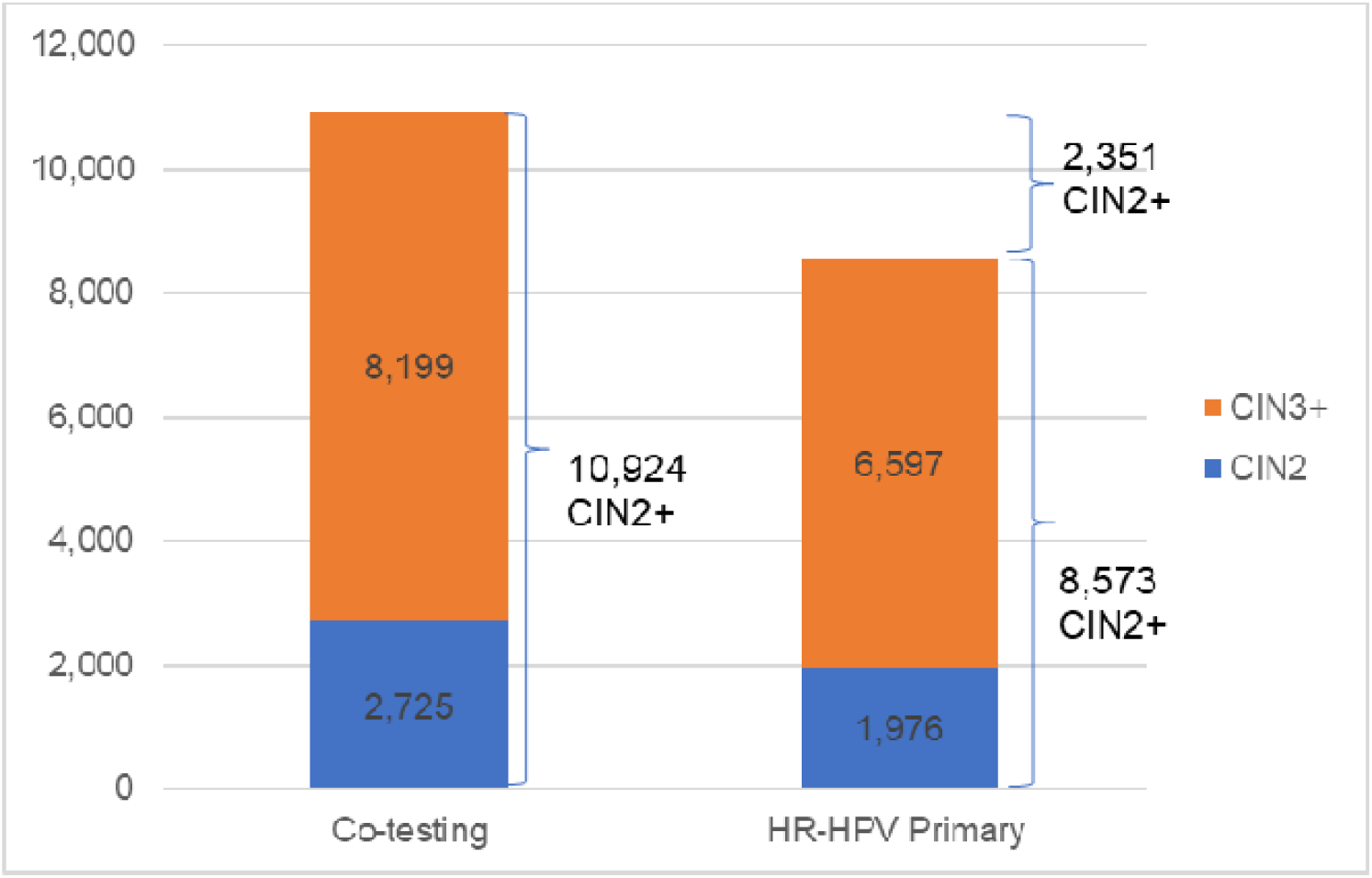
Number of CIN2 and CIN3+ cases diagnosed by screening algorithm.

**Table 2.** Primary outcomes under baseline assumptions for HR-HPV primary and co-testing algorithms for cervical screening (cohort N=577,846)

| Screening Algorithm | Total programme cost (€) | Cost per screen (€) | Number of colposcopies | HR-HPV tests | LBC tests | CIN2+ diagnoses | CIN3+ diagnoses |
| --- | --- | --- | --- | --- | --- | --- | --- |
| Co-testing | 58,775,083 | 101.7 | 28,055 | 645,577 | 652,555 | 10,924 | 8,199 |
| HR-HPV primary | 65,588,573 | 113.5 | 14,882 | 577,846 | 175,535 | 8,573 | 6,597 |

Abbreviations: CIN, Cervical squamous intraepithelial neoplasia; HR-HPV, Human papillomavirus.

**Table 3.** Secondary outcomes under baseline assumptions for HR-HPV primary and co-testing algorithms for cervical screening (cohort N=577,846).

| Screening Algorithm | Colposcopies (€) | HR-HPV tests (€) | LBC tests (€) | CIN 1 treatment (€) | CIN 2 treatment (€) | CIN 3+ treatment (€) |
| --- | --- | --- | --- | --- | --- | --- |
| Co-testing | 1,071,664 | 32,811,275 | 17,825,880 | 1,510,269 | 1,030,170 | 4,525,826 |
| HR-HPV primary | 569,078 | 53,005,826 | 7,233,726 | 810,690 | 747,902 | 3,221,352 |

Abbreviations: CIN – Cervical squamous intraepithelial neoplasia; HR-HPV – Human papillomavirus. CIN 3+ includes cost of treatment of CIN 3 and cervical cancer.

**Table 4.** Average cost (€) and number of colposcopies per CIN 2+ and CIN 3+ case for HR-HPV primary and co-testing algorithms for cervical screening.

| Screening Algorithm | Average cost per CIN2+ case (€) | Average cost per CIN3+ case (€) | Average number of colposcopies per CIN2+ case |
| --- | --- | --- | --- |
| Co-testing | €5,381 | €7,169 | 2.6 |
| HR-HPV primary | €7,650 | €9,942 | 1.7 |

Abbreviations: CIN – Cervical squamous intraepithelial neoplasia; HR-HPV – Human papillomavirus.

### Deterministic sensitivity analysis (DSA)

The impact of varying each input value between its low and high value are presented in **Supplemental Figure 1**. The unit cost of consultation, and of HR-HPV testing and LBC are the main drivers influencing the total cost for both algorithms. Screening coverage was an important driver of the total cost, the total number of colposcopies, LBC, and HR-HPV tests. The probability of CIN3 at 18 months, given a positive HR-HPV test in the HR-HPV primary algorithm or positive results in both tests in the co-testing algorithm at baseline, has an impact on the total number of CIN2+ and CIN3+ cases in both algorithms.

### Probabilistic sensitivity analysis (PSA)

The results of the PSA are shown in **Supplemental Figure 2**. Implementation of the co-testing algorithm led to a greater number of HR-HPV tests, LBC and colposcopies in all iterations compared to the HR-HPV primary algorithm, with a lower total cost in 82.8% of 1,000 iterations. The co-testing algorithm detected more CIN2+ cases in 92.7% and more CIN3+ cases in 87.6% of 1,000 iterations.

### Scenario analysis

Scenario 1 and 2 simulated vaccination coverage of 67% (46,671) and 90% (62,693) of women under 30 (69,659). These changes resulted in a decrease in total costs of 1.4% and 1.9% for HR-HPV primary and 1.6% and 2.2% for co-testing algorithm, respectively, due to fewer positive HR-HPV tests at baseline.

Scenario 3 increased the total cost by 51% compared to using the cost-neutral pricing strategy for co-testing pathway. Total costs for the co-testing pathway were €88.6M compared to the HR-HPV primary pathway total costs of €65.6M. The full results of all scenario analyses are presented in **Supplemental Table 7**.

## Discussion

### Main findings

This modelling study compares two hypothetical algorithms for cervical cancer screening in Belgium. The results indicate that, using the algorithms modelled here, co-testing would detect more cervical precancer and reduce the risks of missing cases and potentially save costs when using a cost-neutral approach for pricing the co-testing package. This comes at an increased use of healthcare resources due to the additional diagnostic tests performed. Uncertainty analyses show that the total costs of both algorithms are sensitive to the reimbursement of HR-HPV and LBC tests. Notably, when applying the same reimbursement rates for these tests, the co-testing algorithm becomes more costly compared to the HR-HPV algorithm. These results can provide useful evidence to help inform decisions on shaping an optimal screening strategy in Belgium.

### Strengths & Limitations

While previous economic modelling studies have compared LBC primary and HR-HPV primary screening (Mendes et al., 2015), research comparing co-testing to HPV primary is limited. This is the first study to compare co-testing to HR-HPV primary testing in cervical screening in Belgium. The model was adapted from previously published cervical screening models (Dombrowski et al., 2022; Weston et al., 2021, 2020), and adapted to the healthcare system in Belgium.

Results from this study align with surveillance data from Germany (co-testing screening) and the Netherlands (HPV primary screening) which provides validation of the model structure and inputs. In the co-testing algorithm, 4.9% of the screening population have colposcopies with 0.5% resulting in the detection of CIN2+ similar to a German co-testing surveillance study reporting approximately 3% ASCUS/AGC or worse results (referral to colposcopy) and 0.5% HSIL or worse (corresponding to CIN2+) in 2021 (Xhaja et al., 2022). In the HR-HPV primary algorithm, 2.6% of the screening population have colposcopies with 1.5% resulting in detection of CIN2+. Likewise in 2021, 2.6% of the screening population in the Netherlands underwent colposcopies and CIN2+ was detected in 1.1% (RIVM, 2021). As with any model, assumptions were made regarding the model structure and inputs. As Belgium does not currently use a co-testing clinical algorithm, a hypothetical co-testing algorithm was adapted from co-testing algorithms in Germany and Luxembourg. There were no data on primary HR-HPV screening or co-testing for Belgium, so data from other trials were used (Rebolj et al., 2016, 2015). This has been deemed an appropriate approach for other studies in the absence of applicable data (EUnetHTA, 2015). Since the model uses HORIZON results for the cobas 4800 HR-HPV assay, it simulates the use of this assay, however, the choice of HR-HPV assay used may impact the outcomes, as different assays have different sensitivity and specificity. The reimbursement for tests may change in the implementation of the national programme in Belgium. Sensitivity analyses found the reimbursement of HPV and LBC tests to be key factors in the total cost and cost per precancer detected. However, the impact of varying these values were assessed in uncertainty analyses and indicated that the main conclusions would not change, lending support for the validity of the modelling work.

The follow-up periods in the algorithms derived from real-world screening algorithms used in neighbouring countries contribute to the difference in number of CIN2+ and CIN3+ detected between the two alternative screening algorithms (shown in **Supplemental Table 5**). The underlying assumption is the probability of detection of cytological abnormalities increases over time, which may overestimate the difference of CIN2+ numbers between two algorithms.

While considering the long-term costs of cancer treatment (co-testing and HR-HPV primary) suggests that co-testing offers improved health and economic outcomes (Felix et al., 2016), the model evaluated the short-term impact of one round of screening including routine screen and follow up and did not include the complexity of disease progression and repeated screening over a lifetime. Taking a simpler approach yields a conservative estimate of the benefits as the true benefits of implementing cervical screening may be underestimated.

### Future work

While it is not necessary to perform a study in Belgium ahead of making decisions to implement a national screening programme, after implementation, the surveillance data from Belgium could be used to validate the results of this study. Moreover, the difference between theoretical and empirical results could be compared, and the model results refined.

Studies in Belgium have found lower rates of HR-HPV infections and cervical abnormalities in the vaccinated population (Arbyn et al., 2016). An increasingly vaccinated population entering the cervical screening programme is likely to have a considerable impact due to an overall reduction in HR-HPV prevalence (Lei et al., 2020), but also a strong relative increase of HPV-negative lesions, and a small absolute increase (Falcaro et al., 2021).

Additional modelling including disease progression, and consideration of new methods to improve cancer detection such as genotype profiling, DNA methylation, artificial intelligence, will be useful in evaluating the optimal screening algorithm and may trigger changing the screening interval or the testing approach (Bedell et al., 2020; Simms et al., 2017).

## Conclusion

These findings support a co-testing cervical screening algorithm in Belgium if the aim is to maximise the number of women with abnormalities found, minimise the risk of missing cases, and minimise the total cost (compared to a HR-HPV strategy) if a cost-neutral discounted co-testing package price was used. The improved detection of pre-cancerous lesions ensures women receive treatment at the earliest opportunity, reducing more severe treatments and mortality.

## Ethics statement

Ethical review and approvals were not required or sought as this paper reports a theoretical health economic evaluation of two cervical screening algorithms and patients were not involved in the evaluation.

## Author Contribution Statement

CD: Methodology, analysis, project administration, interpretation, supervision, writing – review & editing; CB: Writing – review & editing; YM: validation, analysis, visualization, writing – original draft, AM: Conceptualization, methodology, analysis, data curation; AP: Conceptualization, methodology, analysis, data curation; BW: Writing – review & editing; KT: Conceptualization, supervision. SH: supervision, writing – review & editing; EA: Conceptualization, supervision, writing – review & editing; JB: Writing – review & editing; RC: Writing – review & editing; SS Writing – review & editing.

All authors reviewed and approved the final manuscript.

## Data Availability

All data relevant to the study are included in the
online Supplementary Material. No additional data are available.

