## Supplementary material for "An economic evaluation of two cervical screening algorithms in Belgium: HR-HPV primary compared to HR-HPV and liquid-based cytology (LBC) co-testing"

**Contents**

| Description of the HORIZON study | Page 2 |
| --- | --- |
| Table 1. Complete list of input parameters | Page 3 |
| Supplemental Table 2. Deterministic and probabilistic sensitivity analysis cost input parameters range and distribution | Page 9 |
| Supplemental Table 3. Deterministic and probabilistic sensitivity analysis probability input parameters range and distribution | Page 10 |
| Supplemental Table 4. Input parameters used in the scenario analyses | Page 12 |
| Supplemental Table 5. Estimated number of CIN2+ for screening cohort by baseline and follow-up test results | Page 13 |
| Supplemental Table 6. Estimated number of colposcopies, HR-HPV tests and LBC tests for screening cohort by year | Page 14 |
| Supplemental Table 7. Scenario analyses exploring impact of different assumptions on vaccination rate | Page 15 |
| Supplemental Figure 1. Tornado diagrams for deterministic sensitivity analysis | Page 16 |
| Supplemental Figure 2. Probabilistic sensitivity analysis: the difference between co-testing vs HR-HPV primary with 1,000 iterations (co-testing – HR-HPV primary). | Page 18 |
| References | Page 19 |

**S1: Description of the HORIZON study**

The HORIZON study, conducted in Copenhagen Denmark, compared HR-HPV assays in a primary cervical screening population aged 23-65 (Rebolj et al., 2016a, 2015a). HR-HPV tests were run on 6258 residual post-cytology samples collected during routine cervical screening at the Hvidovre University Hospital in Copenhagen between June and August 2011. Within the study, those with abnormal LBC results received repeat testing (LSIL or HPV negative ASCUS-only) or were referred for colposcopy (other abnormal LBC results) in accordance with Danish guidelines at the time. Those with normal LBC and HR-HPV positive results on one or more HR-HPV assays were invited to return for follow up LBC and HPV testing at 18 months. If LBC abnormal or HR-HPV positivity at 18-month follow-up, they were referred for colposcopy. Histology results after colposcopy were reported.

**Supplemental Table 1. Complete list of input parameters**

| **Parameter** | **Age group** | **Screening Strategy** | **Baseline value** |
| --- | --- | --- | --- |
| **Demographic** | | | |
| Screening coverage | Both | Both | 62.6% |
| Total women attending cervical screening | Both | Both | 577,846 |
| - Women aged 25-29 | <30 | Both | 69,659 |
| - Women aged 30-64 | ≥30 | Both | 508,187 |
| Time to recall for a control cytology following a positive co-test at baseline (months) | Both | Both | 6 |
| Time to recall for a repeat co-test following a positive co-test at baseline (months) | Both | Both | 12 |
| Time to recall for a colonoscopy following a positive co-test at baseline (months) | Both | Both | 1 |
| % of HR-HPV negative & LBC positive defined as medium risk at baseline | Both | Co-testing | 0.74 |
| % of colposcopies with biopsy for women who underwent colposcopies | Both | Both | 0.0700 |
| Loss to follow-up for LBC and co-testing for women who had abnormal screening results | Both | Both | 0.3130 |
| Loss to follow-up for colposcopy for women who had abnormal screening results | Both | Both | 0.3130 |
| **Testing and treatment costs** **(all values in €)** | | | |
| HR-HPV test | Both | HR-HPV primary | 67.90 |
| LBC test | Both | HR-HPV primary | 32.41 |
| HR-HPV test plus LBC test | Both | Co-testing^1^ | 54.02 |
| Follow-up LBC test | Both | Co-testing | 32.41 |
| LBC test (control test) | Both | Both | 56.24 |
| Colposcopy with biopsy | Both | Both | 91.40 |
| Colposcopy without biopsy | Both | Both | 34.31 |
| Cost of consultation in which a swab is taken | Both | Both | 23.83 |
| CIN 1 treatment | Both | Both | 284.28 |
| CIN 2 treatment | Both | Both | 378.47 |
| CIN 3 treatment | Both | Both | 488.32 |
| Cervical cancer treatment | Both | Both | 5117.43 |
| Cost annual discount rate | Both | Both | 3.0% |
| Formula for discount based on the recall period | Both | Both | 1.0149 |
| Weighted cost of colposcopy (biopsy & no biopsy) | Both | Both | 38.31 |
| Total cost of HR-HPV testing (including consultation) | Both | Co-testing | 50.84 |
|  | Both | HR-HPV primary | 91.73 |
| **Clinical and diagnostic probabilities (baseline)**^2^ | | | |
| Positive HR-HPV test at baseline | <30 | HR-HPV primary | 0.4061 |
|  | ≥30 | HR-HPV primary | 0.1621 |
| Positive reflex LBC test at baseline | <30 | HR-HPV primary | 0.1676 |
|  | ≥30 | HR-HPV primary | 0.1699 |
| Negative colposcopy (inc. CIN 0) at baseline if HR-HPV test positive at baseline | <30 | HR-HPV primary | 0.2143 |
|  | ≥30 | HR-HPV primary | 0.1897 |
| Negative colposcopy (inc. CIN 0) at baseline if HR-HPV test positive & LBC test positive at baseline | <30 | Co-testing | 0.2143 |
|  | ≥30 | Co-testing | 0.1897 |
| Negative colposcopy (inc. CIN 0) at baseline if HR-HPV test negative & LBC test positive at baseline | <30 | Co-testing | 0.8000 |
|  | ≥30 | Co-testing | 0.6316 |
| Negative HR-HPV test & negative LBC test at baseline | <30 | Co-testing | 0.5876 |
|  | ≥30 | Co-testing | 0.8212 |
| Negative HR-HPV test & positive LBC test at baseline | <30 | Co-testing | 0.7400 |
|  | ≥30 | Co-testing | 0.0043 |
| Positive HR-HPV test & negative LBC test at baseline (for women with medium risk) | <30 | Co-testing | 0.0046 |
|  | ≥30 | Co-testing | 0.0124 |
| Positive HR-HPV test & negative LBC test at baseline | <30 | Co-testing | 0.3380 |
|  | ≥30 | Co-testing | 0.1345 |
| Positive HR-HPV test & positive LBC test at baseline | <30 | Co-testing | 0.0681 |
|  | ≥30 | Co-testing | 0.0275 |
| CIN 1 at baseline if HR-HPV test positive at baseline | <30 | HR-HPV primary | 0.1964 |
|  | ≥30 | HR-HPV primary | 0.1724 |
| CIN 2 at baseline if HR-HPV test positive at baseline | <30 | HR-HPV primary | 0.1429 |
|  | ≥30 | HR-HPV primary | 0.1379 |
| CIN 3 at baseline if HR-HPV test positive at baseline | <30 | HR-HPV primary | 0.4464 |
|  | ≥30 | HR-HPV primary | 0.5000 |
| Cervical cancer at baseline if HR-HPV test positive at baseline | <30 | HR-HPV primary | 0 |
|  | ≥30 | HR-HPV primary | 0 |
| CIN 1 at baseline if HR-HPV test negative & LBC test positive at baseline | <30 | Co-testing | 0 |
|  | ≥30 | Co-testing | 0.2632 |
| CIN 2 at baseline if HR-HPV test negative & LBC test positive at baseline | <30 | Co-testing | 0 |
|  | ≥30 | Co-testing | 0 |
| CIN 3 at baseline if HR-HPV test negative & LBC test positive at baseline | <30 | Co-testing | 0.2000 |
|  | ≥30 | Co-testing | 0.0526 |
| Cervical cancer at baseline if HR-HPV test negative & LBC test positive at baseline | <30 | Co-testing | 0 |
|  | ≥30 | Co-testing | 0.0526 |
| CIN 1 at baseline if HR-HPV test positive & LBC test positive at baseline | <30 | Co-testing | 0.1964 |
|  | ≥30 | Co-testing | 0.1724 |
| CIN 2 at baseline if HR-HPV test positive & LBC test positive at baseline | <30 | Co-testing | 0.1429 |
|  | ≥30 | Co-testing | 0.1379 |
| CIN 3 at baseline if HR-HPV test positive & LBC test positive at baseline | <30 | Co-testing | 0.4464 |
|  | ≥30 | Co-testing | 0.5000 |
| Cervical cancer at baseline if HR-HPV test positive & LBC test positive at baseline | <30 | Co-testing | 0 |
|  | ≥30 | Co-testing | 0 |
| **Clinical and diagnostic probabilities (Follow-up periods)**^2^ | | | |
| Negative colposcopy (inc. CIN 0) at 6 months if HR-HPV test positive & LBC test positive at baseline | <30 | Co-testing | 0.2143 |
|  | ≥30 | Co-testing | 0.1897 |
| Negative colposcopy (inc. CIN 0) at 6 months if HR-HPV test negative & LBC test positive at baseline | <30 | Co-testing | 0.8000 |
|  | ≥30 | Co-testing | 0.6316 |
| Negative colposcopy (inc. CIN 0) at 6 months if medium risk at baseline | <30 | Co-testing | 0.8000 |
|  | ≥30 | Co-testing | 0.6316 |
| Negative colposcopy (inc. CIN 0) at 12 months if HR-HPV test positive & LBC test negative at baseline | <30 | Co-testing | 0.5683 |
|  | ≥30 | Co-testing | 0.6578 |
| Negative colposcopy (inc. CIN 0) at 18 months if HR-HPV test positive & LBC test positive at baseline | <30 | Co-testing | 0.2143 |
|  | ≥30 | Co-testing | 0.1897 |
| Negative colposcopy (inc. CIN 0) at 18 months if HR-HPV test negative & LBC test positive at baseline | <30 | Co-testing | 0.8000 |
|  | ≥30 | Co-testing | 0.6316 |
| Negative colposcopy (inc. CIN 0) at 18 months if medium risk at baseline | <30 | Co-testing | 0.8000 |
|  | ≥30 | Co-testing | 0.6316 |
| Positive control LBC at 6 months if HR-HPV test positive & LBC test positive at baseline | <30 | Co-testing | 0.0371 |
|  | ≥30 | Co-testing | 0.0448 |
| Positive control LBC test at 6 months if medium risk at baseline | <30 | Co-testing | 0.0371 |
|  | ≥30 | Co-testing | 0.0448 |
| Positive control LBC test at 6 months if HR-HPV test negative & LBC test positive at baseline | <30 | Co-testing | 0.0371 |
|  | ≥30 | Co-testing | 0.0448 |
| Positive co-testing at 12 months if HR-HPV test positive & LBC test negative at baseline | <30 | Co-testing | 0.3456 |
|  | ≥30 | Co-testing | 0.2583 |
| Positive co-testing at 18 months if HR-HPV test positive & LBC test positive at baseline | <30 | Co-testing | 0.7470 |
|  | ≥30 | Co-testing | 0.8378 |
| Positive co-testing at 18-month if HR-HPV test negative & LBC test positive at baseline | <30 | Co-testing | 0.8333 |
|  | ≥30 | Co-testing | 0.5349 |
| Positive co-testing at 18 months if medium risk at baseline | <30 | Co-testing | 0.4124 |
|  | ≥30 | Co-testing | 0.1788 |
| Negative colposcopy (inc. CIN 0) at 6 months if HR-HPV test positive & LBC test negative at baseline and control LBC test positive at 6-month follow-up | <30 | HR-HPV primary | 0.3766 |
|  | ≥30 | HR-HPV primary | 0.5079 |
| Negative colposcopy (inc. CIN 0) at 12 months if HR-HPV test positive at baseline | <30 | HR-HPV primary | 0.2143 |
|  | ≥30 | HR-HPV primary | 0.1897 |
| Negative colposcopy (inc. CIN 0) at 18 months if HR-HPV test positive at baseline | <30 | HR-HPV primary | 0.2143 |
|  | ≥30 | HR-HPV primary | 0.1897 |
| Negative colposcopy (inc. CIN 0) at 18 months if HR-HPV test positive & LBC test negative at baseline and control LBC test positive & colposcopy negative at 6-month follow-up | <30 | HR-HPV primary | 0.3766 |
|  | ≥30 | HR-HPV primary | 0.5079 |
| Positive LBC test at 6 months | <30 | HR-HPV primary | 0.0371 |
|  | ≥30 | HR-HPV primary | 0.0448 |
| Positive control LBC test at 12 months if HR-HPV test positive & LBC test positive and colposcopy negative at baseline | <30 | HR-HPV primary | 0.0728 |
|  | ≥30 | HR-HPV primary | 0.0877 |
| Positive control LBC test at 18 months if HR-HPV test positive & LBC test negative at baseline and control LBC test positive and colposcopy negative at 6 months | <30 | HR-HPV primary | 0.1071 |
|  | ≥30 | HR-HPV primary | 0.1286 |
| CIN 1 at 6 months if medium risk at baseline and control LBC test positive at 6-month follow-up | <30 | Co-testing | 0 |
|  | ≥30 | Co-testing | 0.2632 |
| CIN 1 at 6 months if HR-HPV test positive & LBC test positive at baseline and control LBC test positive at 6-month follow-up | <30 | Co-testing | 0.1964 |
|  | ≥30 | Co-testing | 0.1724 |
| CIN 1 at 6 months if HR-HPV test negative & LBC test positive at baseline and control LBC test positive at 6-month follow-up | <30 | Co-testing | 0 |
|  | ≥30 | Co-testing | 0.2632 |
| CIN 1 at 12 months if HR-HPV test positive & LBC test negative at baseline and co-testing positive at 12-month follow-up | <30 | Co-testing | 0.1438 |
|  | ≥30 | Co-testing | 0.2128 |
| CIN 1 at 18 months if HR-HPV test positive & LBC test positive at baseline and co-testing positive at 18-month follow-up | <30 | Co-testing | 0.1964 |
|  | ≥30 | Co-testing | 0.1724 |
| CIN 1 at 18 months if medium risk at baseline and co-testing positive at 6-month follow-up | <30 | Co-testing | 0 |
|  | ≥30 | Co-testing | 0.2632 |
| CIN 1 at 18 months if HR-HPV test negative & LBC test positive at baseline and co-testing positive at 6-month follow-up | <30 | Co-testing | 0 |
|  | ≥30 | Co-testing | 0.2632 |
| CIN 2 at 6 months if medium risk at baseline and control LBC test positive at 6-month follow-up | <30 | Co-testing | 0 |
|  | ≥30 | Co-testing | 0 |
| CIN 2 at 6 months if HR-HPV test positive & LBC test positive at baseline and control LBC test positive at 6-month follow-up | <30 | Co-testing | 0.1429 |
|  | ≥30 | Co-testing | 0.1379 |
| CIN 2 at 6 months if HR-HPV test negative & LBC test positive at baseline and control LBC test positive at 6-month follow-up | <30 | Co-testing | 0 |
|  | ≥30 | Co-testing | 0 |
| CIN 2 at 12 months if HR-HPV test positive & LBC test negative at baseline and co-testing positive at 12-month follow-up | <30 | Co-testing | 0.1252 |
|  | ≥30 | Co-testing | 0.0428 |
| CIN 2 at 18 months if HR-HPV test positive & LBC test positive at baseline and co-testing positive at 18-month follow-up | <30 | Co-testing | 0.1429 |
|  | ≥30 | Co-testing | 0.1379 |
| CIN 2 at 18 months if medium risk at baseline and co-testing positive at 6-month follow-up | <30 | Co-testing | 0 |
|  | ≥30 | Co-testing | 0 |
| CIN 2 at 18 months if HR-HPV test negative & LBC test positive at baseline and co-testing positive at 6-month follow-up | <30 | Co-testing | 0 |
|  | ≥30 | Co-testing | 0 |
| CIN 3 at 6 months if medium risk at baseline and control LBC test positive at 6-month follow-up | <30 | Co-testing | 0.2000 |
|  | ≥30 | Co-testing | 0.0526 |
| CIN 3 at 6 months if HR-HPV test positive & LBC test positive at baseline and control LBC test positive at 6-month follow-up | <30 | Co-testing | 0.4464 |
|  | ≥30 | Co-testing | 0.5000 |
| CIN 3 at 6 months if HR-HPV test negative & LBC test positive at baseline and control LBC test positive at 6-month follow-up | <30 | Co-testing | 0.2000 |
|  | ≥30 | Co-testing | 0.0526 |
| CIN 3 at 12 months if HR-HPV test positive & LBC test negative at baseline and co-testing positive at 12-month follow-up | <30 | Co-testing | 0.1626 |
|  | ≥30 | Co-testing | 0.0866 |
| CIN 3 at 18 months if HR-HPV test positive & LBC test positive at baseline and co-testing positive at 18-month follow-up | <30 | Co-testing | 0.4464 |
|  | ≥30 | Co-testing | 0.5000 |
| CIN 3 at 18 months if medium risk at baseline and co-testing positive at 6-month follow-up | <30 | Co-testing | 0.2000 |
|  | ≥30 | Co-testing | 0.0526 |
| CIN 3 at 18 months if HR-HPV test negative & LBC test positive at baseline and co-testing positive at 6-month follow-up | <30 | Co-testing | 0.2000 |
|  | ≥30 | Co-testing | 0.0526 |
| Cervical cancer at 6 months if medium risk at baseline and control LBC test positive at 6-month follow-up | <30 | Co-testing | 0 |
|  | ≥30 | Co-testing | 0.0526 |
| Cervical cancer at 6 months if HR-HPV test positive & LBC test positive at baseline and control LBC test positive at 6-month follow-up | <30 | Co-testing | 0 |
|  | ≥30 | Co-testing | 0 |
| Cervical cancer at 6 months if HR-HPV test negative & LBC test positive at baseline and control LBC test positive at 6-month follow-up | <30 | Co-testing | 0 |
|  | ≥30 | Co-testing | 0.0526 |
| Cervical cancer at 12 months if HR-HPV test positive & LBC test negative at baseline and co-testing positive at 12-month follow-up | <30 | Co-testing | 0 |
|  | ≥30 | Co-testing | 0 |
| Cervical cancer at 18 months if HR-HPV test positive & LBC test positive at baseline and co-testing positive at 18-month follow-up | <30 | Co-testing | 0 |
|  | ≥30 | Co-testing | 0 |
| Cervical cancer at 18 months if medium risk at baseline and co-testing positive at 6-month follow-up | <30 | Co-testing | 0 |
|  | ≥30 | Co-testing | 0.0526 |
| Cervical cancer at 18 months if HR-HPV test negative & LBC test positive at baseline and co-testing positive at 6-month follow-up | <30 | Co-testing | 0 |
|  | ≥30 | Co-testing | 0.0526 |
| CIN 1 at 6 months if HR-HPV test positive & LBC test negative at baseline and control LBC test positive at 6-month follow-up | <30 | HR-HPV primary | 0.2078 |
|  | ≥30 | HR-HPV primary | 0.3016 |
| CIN 1 at 12 months if HR-HPV test positive at baseline and control LBC test positive at 12-month follow-up | <30 | HR-HPV primary | 0.1964 |
|  | ≥30 | HR-HPV primary | 0.1724 |
| CIN 1 at 18 months if HR-HPV test positive at baseline and control LBC test positive at 18-month follow-up | <30 | HR-HPV primary | 0.1964 |
|  | ≥30 | HR-HPV primary | 0.1724 |
| CIN 1 at 18 months if HR-HPV test positive & LBC test negative at baseline and control LBC test positive & colposcopy negative at 18-month follow-up | <30 | HR-HPV primary | 0.2078 |
|  | ≥30 | HR-HPV primary | 0.3016 |
| CIN 2 at 6 months if HR-HPV test positive & LBC test negative at baseline and control LBC test positive at 6-month follow-up | <30 | HR-HPV primary | 0.1818 |
|  | ≥30 | HR-HPV primary | 0.0635 |
| CIN 2 at 12 months if HR-HPV test positive at baseline and control LBC test positive at 12-month follow-up | <30 | HR-HPV primary | 0.1429 |
|  | ≥30 | HR-HPV primary | 0.1379 |
| CIN 2 at 18 months if HR-HPV test positive at baseline and control LBC test positive at 18-month follow-up | <30 | HR-HPV primary | 0.1429 |
|  | ≥30 | HR-HPV primary | 0.1379 |
| CIN 2 at 18 months if HR-HPV test positive & LBC test negative at baseline and control LBC test positive & colposcopy negative at 18-month follow-up | <30 | HR-HPV primary | 0.1818 |
|  | ≥30 | HR-HPV primary | 0.0635 |
| CIN 3 at 6 months if HR-HPV test positive & LBC test negative at baseline and control LBC test positive at 6-month follow-up | <30 | HR-HPV primary | 0.2338 |
|  | ≥30 | HR-HPV primary | 0.1270 |
| CIN 3 at 12 months if HR-HPV test positive at baseline and control LBC test positive at 12-month follow-up | <30 | HR-HPV primary | 0.4464 |
|  | ≥30 | HR-HPV primary | 0.5000 |
| CIN 3 at 18 months if HR-HPV test positive at baseline and control LBC test positive at 18-month follow-up | <30 | HR-HPV primary | 0.4464 |
|  | ≥30 | HR-HPV primary | 0.5000 |
| CIN 3 at 18 months if HR-HPV test positive & LBC test negative at baseline and control LBC test positive & colposcopy negative at 18-month follow-up | <30 | HR-HPV primary | 0.2338 |
|  | ≥30 | HR-HPV primary | 0.1270 |
| Cervical cancer at 6 months if HR-HPV test positive & LBC test negative at baseline and control LBC test positive at 6-month follow-up | <30 | HR-HPV primary | 0 |
|  | ≥30 | HR-HPV primary | 0 |
| Cervical cancer at 12 months if HR-HPV test positive at baseline and control LBC test positive at 12-month follow-up | <30 | HR-HPV primary | 0 |
|  | ≥30 | HR-HPV primary | 0 |
| Cervical cancer at 18 months if HR-HPV test positive at baseline and control LBC test positive at 18-month follow-up | <30 | HR-HPV primary | 0 |
|  | ≥30 | HR-HPV primary | 0 |
| Cervical cancer at 18 months if HR-HPV test positive & LBC test negative at baseline and control LBC test positive & colposcopy negative at 18-month follow-up | <30 | HR-HPV primary | 0 |
|  | ≥30 | HR-HPV primary | 0 |

Abbreviations: CIN, Cervical squamous intraepithelial neoplasia; DSA, Deterministic sensitivity analysis; LBC, Liquid-based cytology; HR-HPV, High-risk human papillomavirus.

^1^The cost of a co-testing package (HR-HPV and LBC test) performed at a screening interval of 5 years, €54.02, was calculated from the annual cost of an LBC test at a screening interval of 3 years of €32.41.

^2^The transition probabilities are calculated from data reported in the HORIZON studies (Rebolj et al., 2016a, 2015a)*.*

**Supplemental Table 2. Deterministic and probabilistic sensitivity analysis cost input parameters range and distribution (Gamma distribution).**

| **Parameter** | **Baseline value (€)** | **DSA values** | | **PSA - Standard deviation (€)** |
| --- | --- | --- | --- | --- |
|  |  | **Low** | **High** |  |
| Cost of HR-HPV test (HR-HPV primary algorithm) | 67.90 | 47.90 | 87.90 | 10.204082 |
| Cost of LBC (reflex test) | 32.41 | 22.41 | 42.41 | 5.102041 |
| Cost of HR-HPV test and LBC (co-testing algorithm) | 54.02 | 34.02 | 74.02 | 10.204082 |
| Cost of consultation in which a swab is taken | 23.83 | 13.83 | 33.83 | 5.102041 |
| Cost of initial investigation with colposcopy (biopsy) | 91.40 | 71.40 | 111.40 | 10.204082 |
| Cost of initial investigation with colposcopy (no biopsy) | 34.31 | 14.31 | 54.31 | 10.204082 |
| Cost of CIN 1 treatment | 284.28 | 227.42 | 341.14 | 29.008163 |
| Cost of CIN 2 treatment | 378.47 | 302.78 | 454.16 | 38.619388 |
| Cost of CIN 3 treatment | 488.32 | 390.66 | 585.98 | 49.828571 |
| Cost of cervical cancer treatment | 5,117.43 | 4,093.94 | 6,140.92 | 522.186735 |
| Cost annual discount rate | 3.0% | 0.0% | 5.0% | - |

**Supplemental Table 3. Deterministic and probabilistic sensitivity analysis probability input parameters range and distribution (Beta distribution).**

| **Clinical and diagnostic probabilities** | **Age group** | Algorithm | | **Baseline value** | **Low value** | **High value** | **Alpha value** | **Beta Value** |
| --- | --- | --- | --- | --- | --- | --- | --- | --- |
| Screening coverage | Both | Both | | 0.6260 | 0.5524 | 0.7000 | 361,732 | 216,114 |
| Colposcopies with biopsy | Both | Both | | 0.0700 | 0.0677 | 0.0723 | 3197 | 42,475 |
| Loss to follow up for control cytology and co-testing recall | Both | Both | | 0.3130 | 0.2513 | 0.3747 | 67.92 | 149.08 |
| Loss to follow-up for colposcopy | Both | Both | | 0.3130 | 0.2513 | 0.3747 | 67.92 | 149.08 |
| Positive HR-HPV at baseline | <30 | HR-HPV primary | | 0.4061 | 0.3792 | 0.4330 | 519 | 759 |
|  | ≥30 | HR-HPV primary | | 0.1621 | 0.1486 | 0.1756 | 465 | 2404 |
| Negative HR-HPV & positive LBC at baseline | <30 | Co-testing | | 0.7400 | 0.5550 | 0.9250 | 2 | 1276 |
|  | ≥30 | Co-testing | | 0.0043 | 0.0019 | 0.0068 | 12 | 2857 |
| Positive HR-HPV & negative LBC at baseline | <30 | Co-testing | | 0.3380 | 0.3121 | 0.3640 | 432 | 846 |
|  | ≥30 | Co-testing | | 0.1345 | 0.1221 | 0.1470 | 386 | 2483 |
| Positive HR-HPV & positive LBC at baseline | <30 | Co-testing | | 0.0681 | 0.0543 | 0.0819 | 87 | 1191 |
|  | ≥30 | Co-testing | | 0.0275 | 0.0215 | 0.0335 | 79 | 2790 |
| CIN 1 at baseline if HR-HPV positive at baseline | <30 | HR-HPV primary | | 0.0121 | 0.0000 | 0.0407 | 0.67 | 55 |
|  | ≥30 | HR-HPV primary | | 0.0105 | 0.0000 | 0.0366 | 0.61 | 58 |
| CIN 2 at baseline if HR-HPV positive at baseline | <30 | HR-HPV primary | | 0.0085 | 0.0000 | 0.0326 | 0.48 | 56 |
|  | ≥30 | HR-HPV primary | | 0.0082 | 0.0000 | 0.0314 | 0.48 | 58 |
| CIN 3 at baseline if HR-HPV positive at baseline | <30 | HR-HPV primary | | 0.0323 | 0.0000 | 0.0786 | 1.81 | 54 |
|  | ≥30 | HR-HPV primary | | 0.0378 | 0.0000 | 0.0868 | 2.19 | 56 |
| Cancer at baseline if HR-HPV positive at baseline | <30 | HR-HPV primary | | 0.0000 | 0 | 0 | 0 | 0 |
|  | ≥30 | HR-HPV primary | | 0.0000 | 0 | 0 | 0 | 58 |
| **Follow-up period in co-testing** algorithm | | | | | | | | |
|  | **Age group** | **Test result at baseline** | | **Baseline value** | **Low value** | **High value** | **Alpha value** | **Beta Value** |
|  |  | **HR-HPV** | **LBC** |  |  |  |  |  |
| Positive co-test at 18-month follow-up | <30 | Negative | Positive | 0.8333 | 0.5351 | 1.0000 | 5 | 1 |
|  | ≥30 |  |  | 0.5349 | 0.3858 | 0.6840 | 23 | 20 |
|  | <30 | Positive | Negative | 0.4124 | 0.4111 | 0.4136 | 238,282 | 339,564 |
|  | ≥30 |  |  | 0.1788 | 0.1778 | 0.1798 | 103,323 | 474,523 |
|  | <30 | Positive | Positive | 0.7470 | 0.6535 | 0.8405 | 62 | 21 |
|  | ≥30 |  |  | 0.8378 | 0.7539 | 0.9218 | 62 | 12 |
| Positive control cytology at 12-month follow-up | <30 | Positive | Positive | 0.0728 | 0.0169 | 0.1287 | 6 | 77 |
|  | ≥30 |  |  | 0.0877 | 0.0232 | 0.1521 | 6 | 68 |
| Positive control cytology at 18-month follow-up | <30 | Positive | Negative | 0.1071 | 0.0664 | 0.1479 | 24 | 197 |
|  | ≥30 |  |  | 0.1286 | 0.0850 | 0.1721 | 29 | 198 |
| CIN 1 at 18-month follow-up | <30 | Negative | Positive | 0.0000 | 0 | 0 | 0 | 5 |
|  | ≥30 |  |  | 0.2632 | 0.0652 | 0.4612 | 5 | 14 |
|  | <30 | Positive | Positive | 0.1964 | 0.0924 | 0.3005 | 11 | 45 |
|  | ≥30 |  |  | 0.1724 | 0.0752 | 0.2696 | 10 | 48 |
| CIN 1 at 12-month follow-up | <30 | Positive | Negative | 0.1438 | 0.0674 | 0.2203 | 12 | 69 |
|  | ≥30 |  |  | 0.2128 | 0.1117 | 0.3139 | 13 | 50 |
| CIN 2 at 18-month follow-up | <30 | Negative | Positive | 0.0000 | 0 | 0 | 0 | 5 |
|  | ≥30 |  |  | 0.0000 | 0 | 0 | 0 | 19 |
|  | <30 | Positive | Positive | 0.1429 | 0.0512 | 0.2345 | 8 | 48 |
|  | ≥30 |  |  | 0.1379 | 0.0492 | 0.2267 | 8 | 50 |
| CIN 2 at 12-month follow-up | <30 | Positive | Negative | 0.1252 | 0.0531 | 0.1973 | 10 | 71 |
|  | ≥30 |  |  | 0.0428 | 0.0000 | 0.0928 | 3 | 60 |
| CIN 3 at 18-month follow-up | <30 | Negative | Positive | 0.2000 | 0.0000 | 0.5506 | 1 | 4 |
|  | ≥30 |  |  | 0.0526 | 0.0000 | 0.1530 | 1 | 18 |
|  | <30 | Positive | Positive | 0.4464 | 0.3162 | 0.5766 | 25 | 31 |
|  | ≥30 |  |  | 0.5000 | 0.3713 | 0.6287 | 29 | 29 |
| CIN 3 at 12-month follow-up | <30 | Positive | Negative | 0.1626 | 0.0823 | 0.2430 | 13 | 68 |
|  | ≥30 |  |  | 0.0866 | 0.0171 | 0.1560 | 5 | 58 |

**Supplemental Table 4. Input parameters used in the scenario analyses**

| **Parameter** | Algorithm | **Value** |
| --- | --- | --- |
| *Scenario 1: Applying current vaccine coverage rate (67% versus 0% at baseline)* | | |
| HR-HPV vaccine coverage rate | Both | 67.0% |
| Relative risk of HR-HPV vaccine on positive HR-HPV test at baseline | Both | 0.57 |
| *Scenario 2: Applying WHO-targeted vaccine coverage rate (90% versus 0% at baseline)* | | |
| HR-HPV vaccine coverage rate | Both | 90.0% |
| Relative risk of HR-HPV vaccine on positive HR-HPV test at baseline | Both | 0.57 |
| *Scenario 3: Using alternative reimbursement cost (€)* | | |
| HR-HPV test | Co-testing | 67.90 |
|  | HR-HPV primary | 67.90 |
| Liquid-based cytology | Co-testing | 32.41 |
|  | HR-HPV primary | 32.41 |

**Supplemental Table 5. Estimated number of CIN2+ for screening cohort by baseline and follow-up test results**

| **HR-HPV primary** algorithm | | | | |
| --- | --- | --- | --- | --- |
| Baseline test results | HPV+ & LBC+ | HPV+ & LBC- | HPV- (excluded from algorithm) | |
| CIN2+ | 8,052 | 0 | 0 | |
| Follow-up test results | LBC+ | LBC+ | No follow-up | |
| CIN2+ | 63 | 459 | 0 | |
| *Total population* | *18,735* | *91,919* | *467,192* | |
| **Co-testing** algorithm | | | | |
| Baseline test results | HPV+ & LBC+ | HPV+ & LBC- | HPV- & LBC+ | HPV- & LBC- (excluded from algorithm) |
| CIN2+ | 8,052 | 0 | 176 | 0 |
| Follow-up test results | Co-test+ | Co-test+ | Co-test+ | No follow-up |
| CIN2+ | 430 | 2,184 | 82 | 0 |
| *Total population* | *18,735* | *91,919* | *8,938* | *458,254* |

**Supplemental Table 6. Estimated number of colposcopies, HR-HPV tests and LBC tests for screening cohort by year**

| **Algorithm** | **Number of Colposcopies** | | **Number of HR-HPV Tests** | | **Number of LBC Tests** | |
| --- | --- | --- | --- | --- | --- | --- |
|  | **Baseline** | **Follow up** | **Baseline** | **Follow up** | **Baseline** | **Follow up** |
| HR-HPV primary | 12,871 | 2,011 | 577,846 | 0 | 110,654 | 64,880 |
| Co-testing | 14,468 | 13,588 | 577,846 | 67,731 | 577,846 | 74,709 |
| **Total** | 27,339 | 15,598 | 1,155,692 | 67,731 | 688,500 | 139,590 |

**Supplemental Table 7. Scenario analyses exploring impact of different assumptions on vaccination rate**

|  | **Total cost (€)** | **Cost per screened (€)** | **Number of colposcopies** | **Number of HR-HPV Tests** | **Number of LBC tests** | **Number of CIN2+** | **Number of CIN3+** |
| --- | --- | --- | --- | --- | --- | --- | --- |
| ***Baseline*** | | | | | | | |
| Co-testing | 58,775,083 | 101.7 | 28,055 | 645,577 | 652,555 | 10,924 | 8,199 |
| HR-HPV primary | 65,588,573 | 113.5 | 14,882 | 577,846 | 175,535 | 8,573 | 6,597 |
| *Difference** | -6,813,490 | -11.8 | 13,173 | 67,731 | 477,020 | 2,351 | 1,602 |
| *Scenario 1: Applying current vaccine coverage rate (67% versus 0% at baseline)* | | | | | | | |
| Co-testing | 57,810,670 | 100.0 | 25,965 | 640,838 | 647,678 | 10,025 | 7,579 |
| HR-HPV primary | 64,676,947 | 111.9 | 13,818 | 577,846 | 162,620 | 7,967 | 6,148 |
| *Difference** | -6,866,277 | -11.9 | 12,147 | 62,992 | 485,058 | 2,057 | 1,431 |
| *Scenario 2: Applying WHO-targeted vaccine coverage rate (90% versus 0% at baseline)* | | | | | | | |
| Co-testing | 57,479,603 | 99.5 | 25,248 | 639,211 | 646,004 | 9,716 | 7,366 |
| HR-HPV primary | 64,364,001 | 111.4 | 13,453 | 577,846 | 158,187 | 7,759 | 5,994 |
| *Difference** | -6,884,398 | -11.9 | 11,795 | 61,365 | 487,817 | 1,957 | 1,373 |
| *Scenario 3: Using alternative reimbursement cost (€)* | | | | | | | |
| Co-testing | 88,650,193 | 153.4 | 28,055 | 645,577 | 652,555 | 10,924 | 8,199 |
| HR-HPV primary | 65,588,573 | 113.5 | 14,882 | 577,846 | 175,535 | 8,573 | 6,597 |
| *Difference** | 23,061,620 | 39.9 | 13,173 | 67,731 | 477,020 | 2,350 | 1,602 |

* Difference: Results of co-testing algorithm – results of HR-HPV primary algorithm

**Supplemental Figure 1. Tornado diagrams for deterministic sensitivity analysis*: co-testing and HR-HPV primary. A: Total algorithms costs; B: Total number of colposcopies; C: Total number of HR-HPV tests; D: Total number of LBC tests; E: Total number of CIN2+ diagnosis; F: Total number of CIN3+ diagnosis**
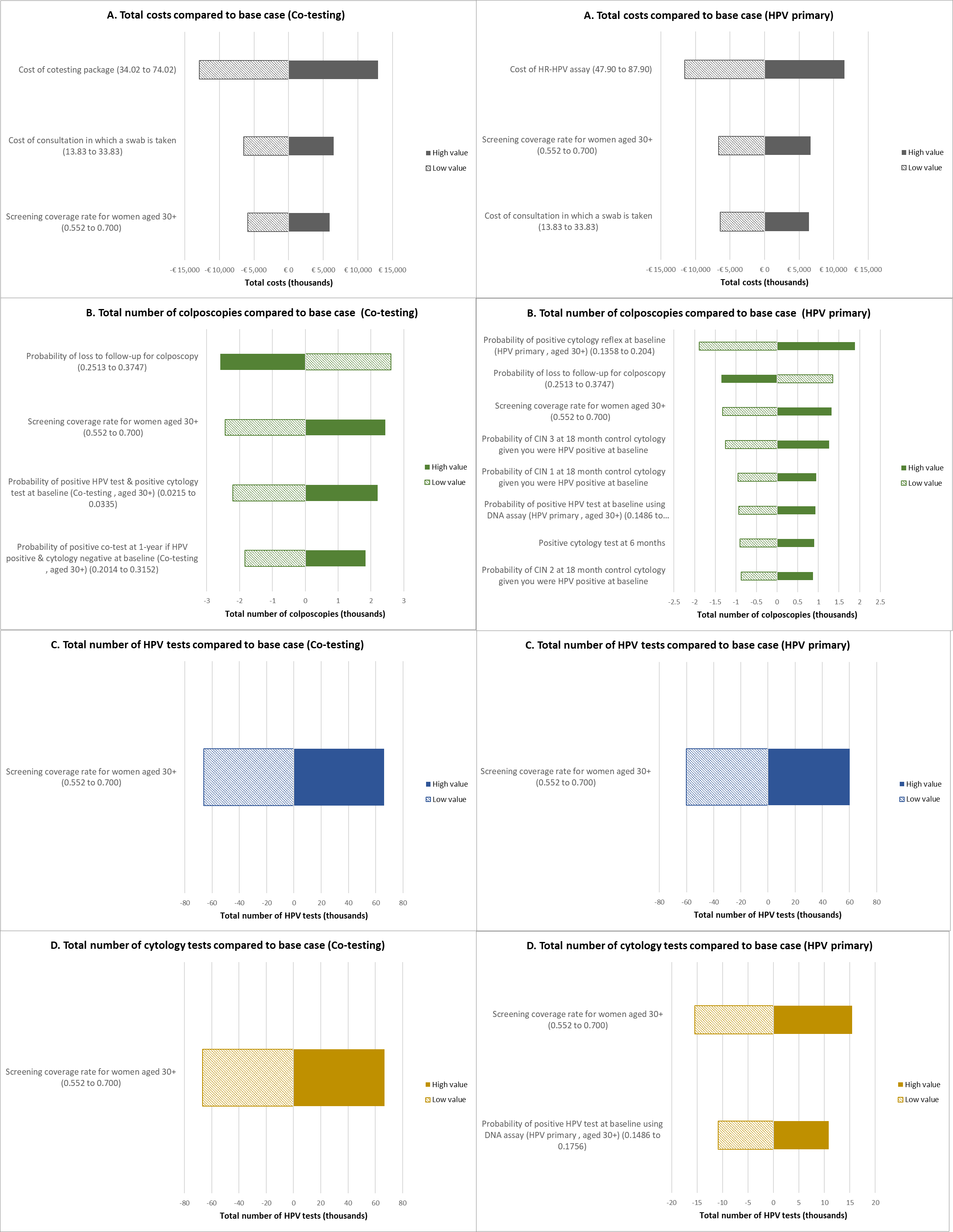

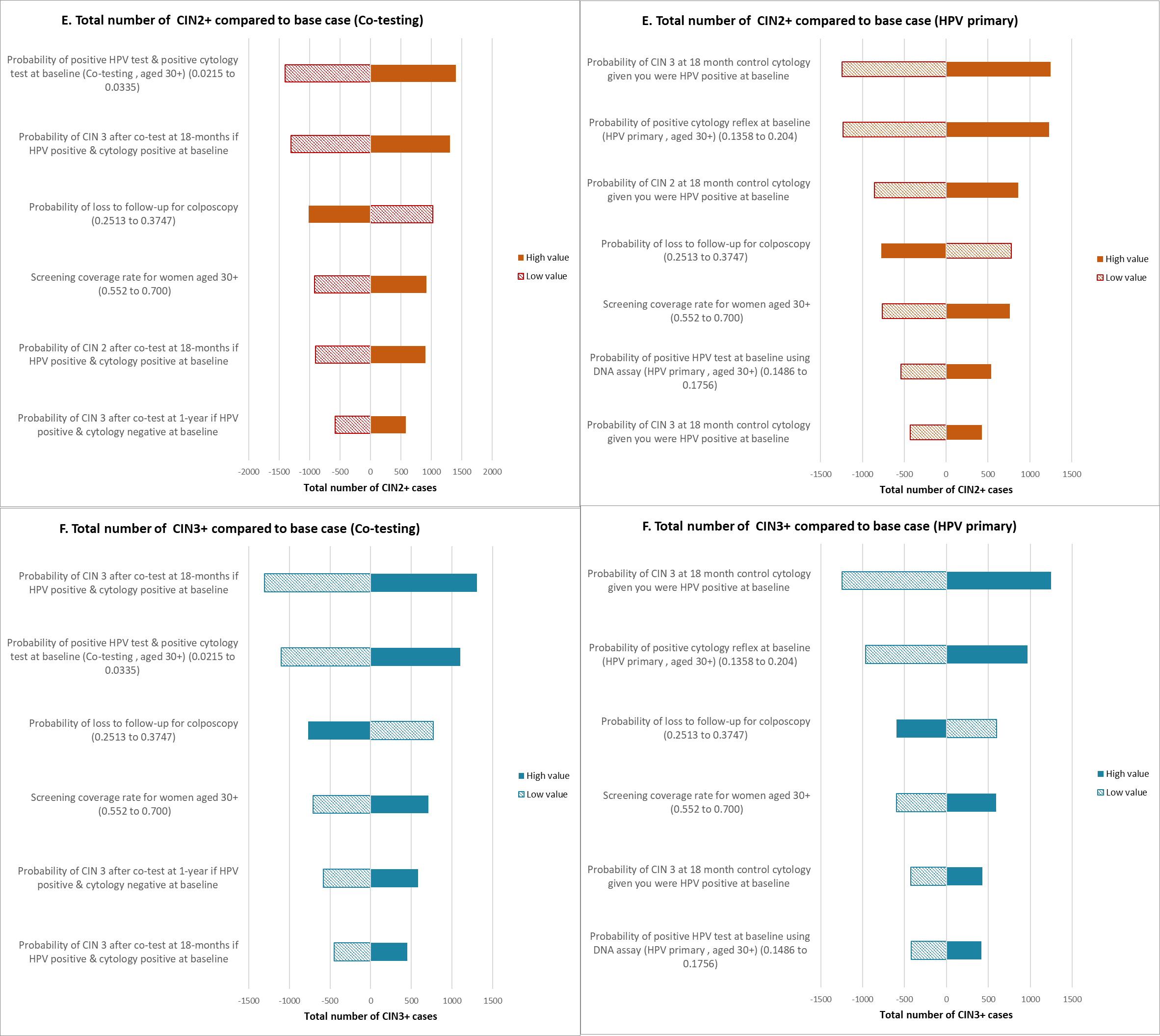

* Parameters that impacted the results by 10% or more are shown in Figure 1 A-F. Solid-filled bars show the outcome using the high value of the input parameter and pattern-filled bars show the outcome using the low value of the input parameter.

**Supplemental Figure 2. Probabilistic sensitivity analysis: the difference between co-testing vs HR-HPV primary with 1,000 iterations (co-testing – HR-HPV primary). A: Difference in costs; B: Difference in number of colposcopies; C: Difference in number of HR-HPV tests; D: Difference in number of LBC tests; E: Difference in CIN2+ diagnosis; F: Difference in CIN3+ diagnosis**
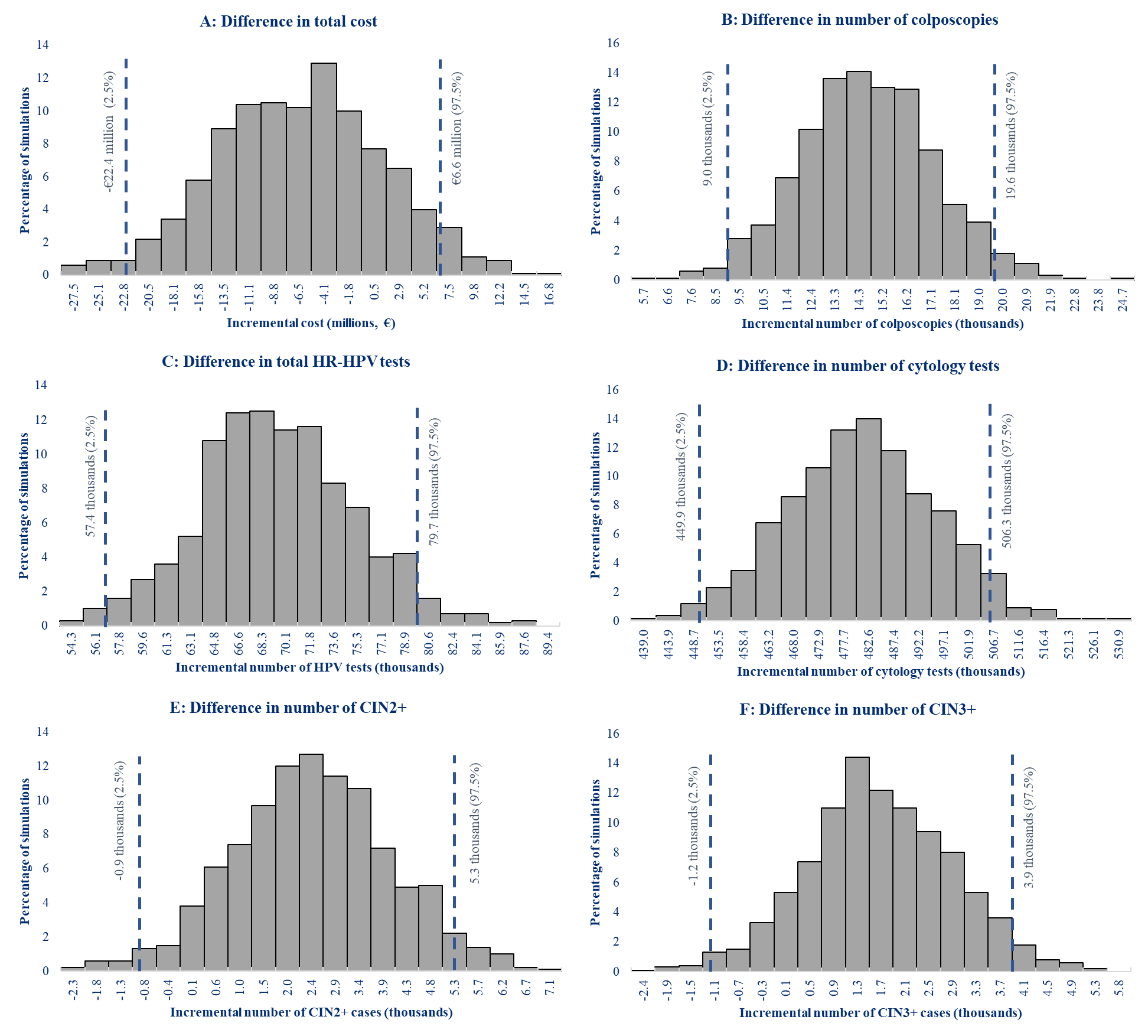

*Note: the negative number in y-axis represents that result in the co-testing algorithmis lower than the result in the HR-HPV primary algorithm. Dashed lines represent 95% Confidence Interval.*
